# Predictors of futile recanalization in patients with large infarct: a post hoc analysis of the ANGEL-ASPECT trial

**DOI:** 10.1101/2023.09.19.23295812

**Authors:** Tingyu Yi, Xiaochuan Huo, Xiao-hui Lin, Mengxing Wang, Yan-Min Wu, Zhi-nan Pan, Xiu-fen Zheng, Ding-lai Lin, Yuesong Pan, Zhongrong Miao, Wen-huo Chen, the ANGEL-ASPECT Investigators

## Abstract

**Background:** Few studies have focused on factors associated with futile recanalization in acute anterior circulation stroke patients with large infarct cores who were treated with modern endovascular therapy (EVT). The aim of this study was to explore the factors associated with futile recanalization in patients with large ischemic strokes.

**Methods:** This is a post hoc analysis of the ANGEL-ASPECT trial. Demographic and clinical characteristics, acute stroke workflow interval times, and imaging characteristics were compared between the futile and meaningful recanalization groups. A favorable outcome was defined as a 90-day mRS score 0-3, successful reperfusion was defined as eTICI 2b, 2c and 3 on final angiogram, and futile recanalization was defined as failure to achieve a favorable outcome despite successful reperfusion. Multivariate analysis was performed to identify the predictors of futile recanalization.

**Result:** One hundred eighty-three patients were included in the final analysis; 91 (49.7%) patients had futile recanalization, and 92 (51.3%) patients had meaningful recanalization. In multivariable logistic regression analysis, older age (age ≥68, OR=3.29, P=0.004), higher NIHSS score (NIHSS ≥ 16, OR=3.33, P=0.003), diabetes (OR=3.23, P=0.017), larger final volume (FIV ≥ 174.7, OR=6.79, P<0.001), postoperative respiratory failure (OR=14.56, P=0.01), and female sex (OR=2.78, P=0.01) were independent predictors of futile recanalization.

**Conclusions:** Futile recanalization occurred in approximately half of acute stroke patients with a large infarct core following endovascular treatment. Old age, high baseline NIHSS score, diabetes mellitus, large FIV and respiratory failure were independent predictors of futile recanalization after endovascular therapy for large ischemic strokes. Stroke-related pneumonia control may improve prognosis.

## Introduction

Endovascular therapy (EVT) is the standard treatment for acute ischemic stroke caused by intracranial large vessel occlusion (LVO)^1–4^; however, the benefit of EVT for patients with a large infarct, defined as an ASPECT score ≤ 5^1,2^ or a baseline infarct volume ≤70 ml, was uncertain until recently, when three breakthrough trials showed the clinical benefit of EVT for large infarcts ^5–7^. According to the results of the ANGEL-ASEPCT trial (Trial of Endovascular Therapy for Acute Ischemic Stroke with Large Infarct, ClinicalTrials.gov number NCT04551664), successful reperfusion (defined as an extended thrombolysis in cerebral infarction (eTICI) score ≥2b) was 83.6%^7^. However, the favorable outcome rate (defined as a modified Rankin scale (mRS) score of 0–3) was 47%; more than half of patients failed to achieve a favorable outcome despite successful recanalization, a phenomenon defined as futile recanalization. The risk factors and incidence of futile recanalization in patients with large infarcts caused by intracranial LVO undergoing EVT have yet to be identified. Thus, in this study, we analyzed data from the ANGEL-ASPECT trial to identify the incidence of and factors predicting futile recanalization.

## Methods

The ANGEL-ASEPCT trial was an investigator-initiated, multicenter, prospective, randomized, open-label trial investigating the outcomes of EVT in Chinese stroke patients with large infarcts caused by the anterior circulation LVO^7^. The study methods and patient eligibility criteria have been reported previously^8^. Patients who were not followed up, did not undergo catheter angiography, or had failed recanalization (mTICI <2b) were excluded from this subgroup analysis.

The baseline information included age, sex, baseline National Institutes of Health Stroke Scale (NIHSS) score, baseline Glasgow Coma Scale (GCS) score, history of hypertension, admission systolic blood pressure (SBP) and diastolic blood pressure, dyslipidemia, diabetes, smoking, atrial fibrillation, previous stroke/transient ischemic attack (TIA), and coronary artery disease. Biochemical tests included serum creatinine, C-reactive protein (CRP), and low-density lipoprotein (LDL) levels.

The cause of stroke included cardioembolism, large-artery atherosclerosis (LAA), undetermined causes or other determined causes. The symptom onset to treatment time, reperfusion time, whether intravenous thrombolysis was performed, time of endovascular procedure initiation, and recanalization time were also recorded.

All patients underwent head CT on admission; the baseline infarct range was assessed by the Alberta Stroke Program Early CT Score (ASPECTS) with noncontrast CT; and the location of the intracranial artery occlusion was identified on digital subtraction angiography (DSA), which included the internal carotid artery (ICA), proximal M1, distal M1 and M2, and ipsilateral extracranial ICA. Perfusion maps and ischemic core volumes (CTP volumes) were determined using RAPID software, version 5.0.4 (iSchemaView). A time to maximum of the residue function [Tmax] exceeding 6 seconds (Tmax> 6 s) was defined as hypoperfusion, and ischemic core volumes were those with a relative CBF (rCBF) < 30%. The mismatch volume was defined as the volume with Tmax> 6 s minus the volume with an rCBF < 30%.

The eTICI score assessed on the final angiogram indicated successful reperfusion, which was graded as 2b56%, 2b67%, 2c or 33. Perioperative medication, operations and complications, including thrombectomy attempts, distal embolism, embolization into a new territory, arterial dissection, contrast leakage, status epilepticus and respiratory failure post-operation, were also recorded. The final infarct volume (FIV) was assessed on follow-up MR on days 5–7 using an automated algorithm.

## Statistical analysis

In the analysis of clinical and radiological outcomes, patients were dichotomized into the meaningful recanalization group (defined as a 90-day mRS score ≤3) and the futile recanalization group (defined as a 90-day mRS score >3). Continuous data are summarized as the median and interquartile range (IQR) or mean and standard deviation (SD), and between-group differences were assessed by *t* tests and Mann‒Whitney U tests, respectively. Categorical variables are described as proportions and were compared using the χ2 test. Variables with a *p* value<0.05 were entered into multivariate logistic regression analysis with a stepwise likelihood ratio model. Data are presented as the odds ratio (OR) and its 95% confidence interval (95% CI). Receiver operating characteristic (ROC) analysis was used to explore the cutoff value of predictive factors. All analyses were performed using SAS version 9.4 (SAS Institute Inc., Cary, NC), with a significance level of *p*<0.05 (two-sided).

## Results

Between October 2, 2020, and May 18, 2022, a total of 1504 patients underwent screening at 46 centers, of whom 456 (30.3%) were enrolled in the ANGEL-ASPECT trial. Of these, 231 were randomly assigned to the endovascular therapy group and 225 to the medical management group. The legal representatives of 1 patient who had been assigned to the thrombectomy group withdrew the patient’s consent, so 230 patients in the thrombectomy group were included in the full-analysis population. Eleven patients were excluded after adjudication, and 219 patients were finally included^7^. Successful reperfusion was achieved in 183 (83.6%) patients, who were included in the analysis (**Fig. 1**).

**Figure 1.**
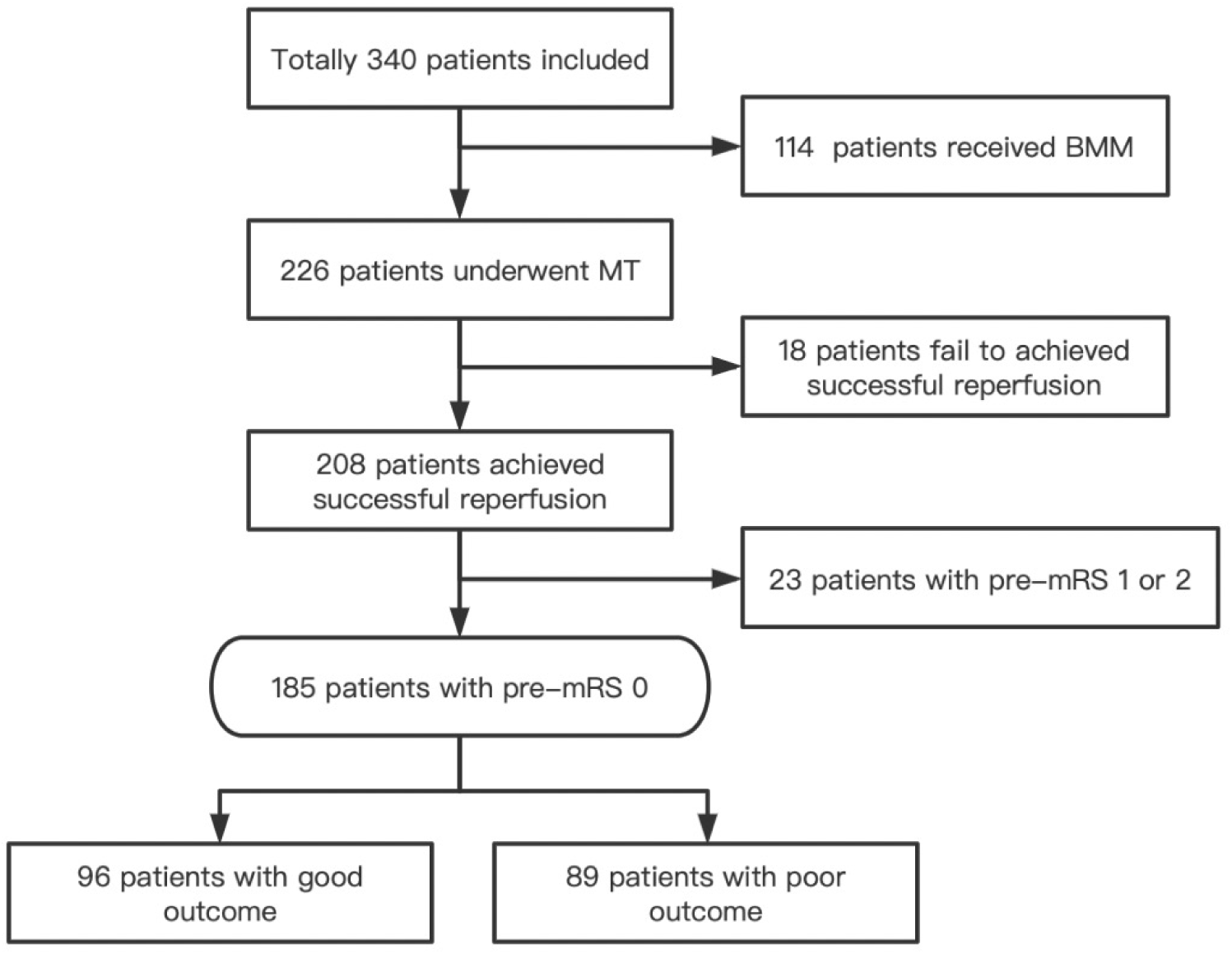
Study Flow Chart. A total of 456 (30.3%) patients were enrolled in the ANGEL-ASPECT trial. Of these, 231 were randomly assigned to the endovascular therapy group and 225 to the medical management group. Successful reperfusion was achieved in 183 (83.6%) patients, who were included in the analysis.

Approximately 49.7% (91/183) of patients achieved futile recanalization, 38.4% (35/91) of them died, while 50.2% (92/183) achieved meaningful recanalization. The baseline characteristics of the patients are listed in **Table 1**. Compared with the patients in the meaningful recanalization group, the patients in the futile recanalization group were significantly older (mean 70 vs. 64 years, *P*<0.001); had significantly higher baseline NIHSS scores (median 18 vs. 14, *P*<0.001), higher baseline systolic pressures (mean 150 vs. 143, *P*=0.005), and larger baseline and final infarct core volumes (median; baseline: 65 vs. 48.50, *P*=0.07; final: 148.25 vs. 97.34, *P*<0.001); and were more likely to be female (48.2% vs. 28.3%, *P*<0.001), to have diabetes (28.6% vs. 12.0%, P<0.006), and to have cardioembolism (60.4% vs. 41.3%, *P*=0.049), less likely to be current smokers (18.7% vs. 35.9%, *P*=0.01), more likely to have symptomatic intracranial hemorrhage (sICH) (10.99% vs. 1.09%, *P*=0.02), and likely to have more respiratory failure (14.3% vs. 1.1%, *P*=0.01).

**Table 1.** Baseline characteristics of patients with futile and meaningful recanalizations.

|  | All Patients (n=183) | Futile recanalization<br>(n=91) | Meaningful recanalization<br>(n=92) | *P* value |
| --- | --- | --- | --- | --- |
| Age (y), mean | 67±9 | 70±8 | 64±9 | <0.001 |
| <68, no. (%) | 84(45.9) | 28(30.8) | 56(60.9) |  |
| ≥68, no. (%) | 99(54.1) | 63(69.2) | 36(39.1) | <0.001 |
| Female sex, no.(%) | 79(43.2) | 53(48.2) | 26(28.3) | <0.001 |
| NIHSS score at admission, median (IQR) | 16(13-20) | 18(14-21) | 14(12-19) | <0.001 |
| < 16, no. (%) | 82(44.8) | 26(28.6) | 56(60.9) |  |
| ≥16, no. (%) | 101(55.2) | 65(71.4) | 36(39.1) | <0.001 |
| Baseline GCS score, no.(%) | 12(9-15) | 11(8-14) | 13(10-15) | 0.047 |
| Medical history |  |  |  |  |
| Hypertension, no.(%) | 105(57.4) | 52(57.1) | 53(57.6) | 0.95 |
| Diabetes, no.(%) | 37(20.2) | 26(28.6) | 11(12.0) | 0.005 |
| Hyperlipidemia, no.(%) | 7(3.8) | 4(4.4) | 3(3.3) | 0.69 |
| Atrial fibrillation, no.(%) | 47(25.7) | 28(30.8) | 19(20.7) | 0.12 |
| Current smoking, no.(%) | 50(27.3) | 17(18.7) | 33(35.9) | 0.01 |
| Blood pressure (mmHg), mean (SD) |  |  |  |  |
| Systolic | 146±25 | 150±25 | 143±23 | 0.06 |
| Diastolic | 85(73-96) | 82(73-96) | 85(74.50-96) | 0.93 |
| Intravenous thrombolysis, no. (%) | 53(29.0) | 24(26.4) | 29(31.5) | 0.44 |
| Awake with stroke symptoms, no. (%) | 58(31.7) | 27(29.7) | 31(33.7) | 0.56 |
| Interval between stroke onset and hospital arrival (min), median (IQR) | 330(192-611) | 330(212-598) | 329.50(173.50-671) | 0.83 |
| Distribution, no. (%) |  |  |  | 0.37 |
| < 4.5 hr | 71(38.8) | 35(38.5) | 36(39.1) |  |
| 4.5 to <6.0 hr | 28(15.3) | 14(15.4) | 14(15.2) |  |
| 6.0 to <12.0 hr | 48(26.2) | 28(30.8) | 20(21.7) |  |
| 12.0 to <24.0 hr | 36(19.7) | 14(15.4) | 22(23.9) |  |
| Stroke classification, no. (%) |  |  |  | 0.04 |
| Atherothrombotic | 46(25.1) | 19(20.9) | 27(29.4) |  |
| Cardioembolic | 93(50.8) | 55(60.4) | 38(41.3) |  |
| Other or unknown | 44(24.0) | 17(18.7) | 27(29.4) |  |
| Laboratory tests |  |  |  |  |
| Creatinine (mg/dL), median (IQR) | 69.9(57-86) | 66.7(53.30-85) | 71.3(59.8-86.0) | 0.42 |
| CRP (mg/dL), median (IQR) | 2.1(1-4.3) | 3.2(1.30-5.44) | 1.7(0.9-3.2) | 0.20 |
| LDL (mmol/L), median (IQR) | 2.76(2.02-3.26) | 2.43(1.67-3.17) | 2.84(2.50-3.36) | 0.15 |
| ASPECTS value based on CT§, median (IQR) | 3(3-4) | 3(3-4) | 3(3-4) | 0.29 |
| 0 | 4(2.19) | 1(1.1) | 3(3.3) |  |
| 1 | 10(5.5) | 8(8.8) | 2(2.2) |  |
| 2 | 9(5.0) | 4(4.4) | 5(5.4) |  |
| 3 | 80(43.7) | 42(46.2) | 38(41.3) |  |
| 4 | 50(27.3) | 24(26.4) | 26(28.3) |  |
| 5 | 30(16.4) | 12(13.2) | 18(19.6) |  |
| Occlusion site, no. (%) |  |  |  | 0.34 |
| ICA | 63(34.4) | 35(38.5) | 28(30.4) |  |
| M1 segment | 119(65.0) | 56(61.5) | 63(68.5) |  |
| M2 segment | 1(0.6) | 0(0.00) | 1(1.1) |  |
| Ipsilateral extracranial ICA occlusion, no. (%) | 31(16.9) | 15(16.5) | 16(17.4) | 0.87 |
| Baseline median infarct-core volume (IQR) | 60(28-85) | 65(31-89) | 48.5(24-78) | 0.07 |
| Baseline mismatch ratio, median (IQR) | 3.05(2.1-5.) | 2.80(1.9-5.0) | 3.30(2.3-6.6) | 0.70 |
| Type of anesthesia during the procedure |  |  |  | 0.44 |
| Local anesthesia | 83(45.36) | 40(43.96) | 43(46.74) |  |
| General anesthesia | 69(37.7) | 38(41.76) | 31(33.70) |  |
| Local anesthesia+conscious sedation (CS). | 31(16.94) | 13(14.29) | 18(19.57) |  |
| Tirofiban use, no. (%) | 89(48.63) | 40(43.96) | 49(53.26) | 0.21 |
| Heparin used, no. (%) | 1(0.55) | 0(0.00) | 1(1.10) | 0.31 |
| Onset-to-reperfusion time (min), median (IQR) | 552(389-835) | 552(389-783) | 554(389.50-924) | 0.58 |
| FPE, no. (%) | 33(18.03) | 15(16.48) | 18(19.57) | 0.59 |
| Thrombectomy attempts, median (IQR) | 2(1-2) | 2(1-3) | 1(1-2) | 0.25 |
| Reperfusion degree, no. (%) |  |  |  | 0.56 |
| eTICI 2b50 | 49(26.8) | 25(27.5) | 24(26.1) |  |
| eTICI 2b 67 | 49(26.8) | 28(30.8) | 21(22.8) |  |
| eTICI 2C | 42(23.0) | 18(19.8) | 24(26.1) |  |
| eTICI 3 | 43(23.5) | 20(22.0) | 23(25.0) |  |
| Complications during the procedure, no. (%) |  |  |  |  |
| Distal embolism | 22(12.1) | 10(11.1) | 12(13.0) | 0.69 |
| New territory embolism | 8(4.4) | 5(5.6) | 3(3.3) | 0.45 |
| Vessel dissection | 1(0.6) | 0(0.0) | 1(1.1) | 0.32 |
| Contrast leakage | 47(25.8) | 22(24.4) | 25(27.2) | 0.67 |
| sICH according to Heidelberg criteria within 48 hr, no. (%) | 11(6.0) | 10(11.0) | 1(1.1) | 0.005 |
| Final infarct volume (mL), median (IQR) | 121.08(62.3-186.5) | 148.25(81.2-243.5) | 97.34(43.1-156.4) | <0.001 |
| < 174.7 ml, no. (%) | 133(73.1) | 51(56.7) | 82(89.2) |  |
| ≥174.7 ml, no. (%) | 49(26.9) | 39(43.3) | 10(10.9) | <0.001 |
| Postoperative complication no.(%) |  |  |  |  |
| Status epilepticus | 1(0.6) | 1(1.1) | 0(0.0) | 0.31 |
| Respiratory failure | 14(7.7) | 13(14.3) | 1(1.1) | 0.01 |

Approximately 22.4% (41/183) of patients were aged ≥75 years, and only approximately 26.8% (11/41) of them had meaningful recanalization. Approximately 67.1% (53/79) of female patients, 65.4% (63/99) of patients aged ≥68 years, 64.4% (65/101) with an NIHSS score ≥16, 70.3% (26/37) of patients with diabetes, 59.1% (55/93) of patients with cardioembolism, 79.6% (39/49) of patients with a final infarct volume ≥174.7 ml, and 92.9% (13/14) of patients with respiratory failure had futile recanalization. Approximately 6.0% (11/183) of patients had sICH; of these, 90.9% (10/11) had futile recanalization, and 54.5% (6/11) died.

ROC analysis was used to predict a functional independence outcome by age, NIHSS score and final infarct volume. The analysis revealed cutoff values of age, NIHSS score and final infarct volume of 68 years, 16 and 174.7 ml, respectively.

In multivariable logistic regression analysis (table 2), the following factors were associated with futile recanalization: older age (age ≥68 vs. <68, OR=3.29, 95% CI 1.48 to 7.34, *P*=0.004), female sex (OR=2.78, 95% CI 1.28 to 6.25, *P*=0.01), higher NIHSS score (NIHSS ≥16 vs. <16, OR=3.33, 95% CI 1.53 to 7.27, *P*=0.003), diabetes (OR=3.23, 95% CI 1.23 to 8.5, *P*=0.017), larger final volume (FIV ≥ 174.7 ml vs. <174.7 ml, OR=6.79, 95% CI 2.61 to 17.69, *P*<0.001), and postoperative respiratory failure (OR=14.56, 95% CI 1.71 to 124.16, *P*=0.01).

## Discussion

To our knowledge, this is the first study to focus on futile recanalization in acute ischemic stroke patients with large infarcts who underwent thrombectomy. Our study showed that approximately half of the patients had futile recanalization, of whom 38.4% died.

Our study showed that factors including older age, higher NIHSS score, diabetes, larger final volume, respiratory failure post treatment and female sex were associated with futile recanalization, which were also predictors of futile recanalization in patients without large infarcts^9^.

In this study, older age was a strong predictor of future recanalization, in line with previous studies^10,11^ conducted in both the modern and the earlier endovascular era. Advanced age is often associated with more cardiovascular risk factors that influence the myogenic tone of blood vessels and self-regulating ability of arteries^12^ and attenuate endothelial function, which may lead to poor collateral circulation^13,14^, increased vessel tortuosity, and increased leukoaraiosis. Poor collateral circulation was associated with an increased risk of conversion of penumbra tissue into infarction and stroke severity^13,15^. Increased vessel tortuosity may increase the difficulties of endovascular procedures, which may lead to increased thrombectomy attempts and a prolonged puncture-to-reperfusion time. All these factors are related to poor outcomes. Age was a negative predictor of clinical outcomes in both patients with and without a large infarct, but the predictive cutoff value for age in patients with a large infarct core was different from that in patients without an infarct core. The ANGEL registry study showed a twofold higher risk of poor clinical outcomes in patients aged > 74 years^10^, but our study showed that in patients with a large infarct core, the risk of poor clinical outcome was threefold higher in patients aged > 68, even using a loose definition of good clinical outcome (mRS score ≥3). A study by Osama O showed that none of the patients with ASPECTS of 0 to 5 and aged >75 years had functional independence at 90 days (mRS score 0-2), but our study showed that approximately one-quarter of patients aged >75 years and ASPECTS of 3-5 had independent ambulation at 90 days (mRS score 0-3). The discrepancy between two studies may be due to different clinical outcome definition. Our study findings also suggest that even advanced patients can benefit from reperfusion therapy despite the low rate.

Our study showed that nearly 70% of female patients had futile recanalization, and female sex was an independent predictor of futile recanalization. The influence of sex on the clinical outcomes of patients who receive EVT is debatable. Some studies have shown no difference between sexes^16,17^, but others have shown that female patients were more likely to have poor outcomes^18,19,20^. Estrogen and sex chromosomes could influence stroke severity and clinical outcomes^21^. Estrogen was related to a smaller infarct volume, and the second X chromosome was related to larger infarct volumes and severe stroke damage^21^. At the reproductive stage, the impact of estrogen is dominant; however, at the reproductive senescence phase, the impact of the second X chromosome is dominant^21^. The average age of the first stroke attack among women was 72.9 years old^22^, which typically represents the reproductive senescence phase, so the impact of the second X chromosome would be dominant. Furthermore, women older than 75 years comprise approximately 60% of the population with atrial fibrillation (AF), which was related to a higher risk of severe stroke due to embolic events^23,24^. Other factors influencing the clinical outcome among female patients are listed as follows. First, due to very restricted social activities and relative ignorance of their health because of the Confucian understanding of women^25,26^ and the higher occurrence of aphasia and reduced level of onsciousness at presentation, women older than 75 years often arrive late to the hospital. Second, at the age when stroke first presents, women are often living alone and therefore have less social support, which contributes to their institutionalization at discharge^22,26^. Third, women are more likely to develop poststroke depression, which is also related to a negative impact on recovery^21^. Fourth, frailty and musculoskeletal comorbidities such as arthritis and osteoporosis are common in female patients^21^.

Diabetes mellitus was also independently associated with futile recanalization and observed in 70% of patients in that group. Dysglycemia can lead to harmful effects in arterial ischemic stroke (AIS) patients, including altered blood‒brain barrier permeability, impaired cerebrovascular reactivity in the microvasculature, increased lactic acid production in ischemic tissues, antifibrinolytic effects and increased vulnerability to reperfusion injury, all of which facilitate infarct growth, brain edema and hemorrhagic transformation^27,28^.

Approximately 6% of patients suffered from sICH, all except one had poor outcomes, and more than half of the patients died, which was in line with a previous study^29,30^. sICH was associated with futile recanalization in univariable analysis, but this association was lost in multivariable analysis. The reason may be due to the small proportion of patients with sICH. Of note, sICH occurrence was moderate in our study and comparable with that in patients without large infarcts^31^. The low sICH occurrence may be due to the highly developed EVT skills and equipment; the median number of thrombectomy attempts was low, and thus occurrence of contrast leakage was also lower than that in a previous study^32^.

Our study showed that FIV was associated with poor clinical outcome and is an independent predictor of futile recanalization, which is consistent with previous studies^33,34,35,36^. However, the optimal cutoff FIV value is controversial. Syed F’s study showed that the optimal FIV cutoff value as a predictor of a favorable outcome was 40 ml, while Albert J found that an FIV of approximately 50 ml demonstrated the greatest accuracy in distinguishing good versus poor outcomes, and an FIV of approximately 90 ml was highly specific for a poor outcome^33,37^. Studies have also shown that the optimal infarct volume distinguishing good from poor outcomes is also related to age^38^ and infarct location^39^. In our study, the cutoff value of FIV predicting a favorable outcome was larger than that in above mentioned studies, which may be due to the different definitions of a favorable outcome and different study samples. Another study showed that the optimal catastrophic clinical outcome (defined as an mRS score 5-6) predictive cutoff volume was 181 mL on apparent diffusion coefficient maps in patients with large middle cerebral artery infarcts who underwent decompressive hemicraniectomy^36^.

Respiratory failure was not common in patients with large infarcts, but once it occurred, almost all patients had unfavorable outcomes, and respiratory failure was related to futile recanalization. Respiratory failure may be related to stroke-associated pneumonia (SAP) and a large infarct core^40^. SAP is one of most common post-stroke infections, occurring with an overall incidence ranging from 5% to 26%^41,42^. The mechanisms by which pneumonia occurs are multifactorial. First, stroke induces local inflammation in situ and systemic immunodepression, which is detected within a few hours after the induction of ischemia and lasts for several weeks^43^. Catecholamine-mediated defects in early lymphocyte activation are the key factor in the impaired antibacterial immune response after stroke^43^. Second, aspiration is related to impaired consciousness and dysphagia^44^. Third, additional risk factors for pneumonia include severe stroke, immobility, weak cough, and placement of a nasogastric feeding tube^40^. Pneumonia is associated with prolonged hospital stay, early and long-term mortality and functional outcome^41^.

Most of the factors associated with futile reperfusion mentioned above, including age, NIHSS score, sex, diabetes mellitus, and FIV (but not respiratory failure) are unmodifiable. So, control respiratory failure may be very important in patients with large infarct and underwent EVT. Since pneumonia is associated with respiratory failure, so strong steps including usage of antibiotic drug, arrangement of anti-aspiration, care of respiratory tract should be taken to control of stroke-related pneumonia, which also hints the importance of postoperative respiratory airway management in patients with large infarcts.

The main strengths of our study were that the dataset was acquired through a nationwide multicentric randomized controlled trial of consecutive thrombectomy procedures and that a systematic, independent 90-day follow-up with adjudication of clinical outcomes was performed. However, there are some limitations. First, although the data were prospectively registered, all data in this subgroup analysis were assessed retrospectively. Second, the sample size was not large enough to explore other variables that are associated with futile recanalization.

## Conclusion

In our post hoc analysis, we observed that futile recanalization occurred in approximately half of patients with large infarcts following EVT. Old age, a high baseline NIHSS score, diabetes mellitus, a large FIV and respiratory failure were independent predictors of futile recanalization for stroke patients with large infarct cores caused by anterior circulation intracranial LVO and underwent EVT. Furthermore, stroke-related pneumonia controlling may reduce respiratory failure occurrence and improve the prognosis of such group’s patients.

## Data Availability

The data that support the findings of this study are available on request from the corresponding author.

## Acknowledgements

We thank the team of American Journal Experts for editing the English text of a draft of this manuscript (https://secure.aje.com).

Search Terms: All Neurology, Cerebral Vascular, Acute Ischemic Stroke, Large infarct

Publication History: NA

